# Prevalence and clinical features of 2019 novel coronavirus disease (COVID-19) in the Fever Clinic of a teaching hospital in Beijing: a single-center, retrospective study

**DOI:** 10.1101/2020.02.25.20027763

**Authors:** Ying Liang, Jingjin Liang, Qingtao Zhou, Xiaoguang Li, Fei Lin, Zhonghua Deng, Biying Zhang, Lu Li, Xiaohua Wang, Hong Zhu, Qingbian Ma, Xiaomei Tong, Jie Xu, Yongchang Sun

## Abstract

**Background:** With the spread of COVID-19 from Wuhan, Hubei Province to other areas of the country, medical staff in Fever Clinics faced the challenge of identifying suspected cases among febrile patients with acute respiratory infections. We aimed to describe the prevalence and clinical features of COVID-19 as compared to pneumonias of other etiologies in a Fever Clinic in Beijing.

**Methods:** In this single-center, retrospective study, 342 cases of pneumonia were diagnosed in Fever Clinic in Peking University Third Hospital between January 21 to February 15, 2020. From these patients, 88 were reviewed by panel discussion as possible or probable cases of COVID-19, and received 2019-nCoV detection by RT-PCR. COVID-19 was confirmed by positive 2019-nCoV in 19 cases, and by epidemiological, clinical and CT features in 2 cases (the COVID-19 Group, n=21), while the remaining 67 cases served as the non-COVID-19 group. Demographic and epidemiological data, symptoms, laboratory and lung CT findings were collected, and compared between the two groups.

**Findings:** The prevalence of COVID-19 in all pneumonia patients during the study period was 6.14% (21/342). Compared with the non-COVID-19 group, more patients with COVID-19 had an identified epidemiological history (90.5% versus 32.8%, *P*<0.001). The COVID-19 group had lower WBC [5.19×10^9^/L (±1.47) versus 7.21×10^9^/L (±2.94), *P*<0.001] and neutrophil counts [3.39×10^9^/L (±1.48) versus 5.38×10^9^/L (±2.85), *P*<0.001] in peripheral blood. However, the count of lymphocytes was not different. On lung CT scans, involvement of 4 or more lobes was more common in the COVID-19 group (45% versus 16.4%, *P*=0.008).

**Interpretation:** In the period of COVID-19 epidemic outside Hubei Province, the prevalence of COVID-19 in patients with pneumonia visiting our Fever Clinic in Beijing was 6.14%. Epidemiological evidence was important for prompt case finding, and lower blood WBC and neutrophil counts may be useful for differentiation from pneumonia of other etiologies.

**Funding:** None.

## Introduction

The first cluster of cases of pneumonia with unknown origin emerged in the city of Wuhan, Hubei Province, China, in early December, 2019 ^1,2^. A novel coronavirus had been isolated from these patients, and subsequently named as 2019 novel coronavirus (2019-nCoV) on January 12, 2020 ^3^, and later as SARS-CoV-2. The initially confirmed cases were localized in Wuhan and mostly associated with Huanan Seafood Wholesale market in the city ^2^, but soon the disease spread rapidly to other areas of the country. On January 19, 2020, two cases in Beijing and one case in Guangdong Province were reported ^4^, and subsequently more cases were reported elsewhere ^5-8^.

Since the outbreak in Wuhan, as a quick response, Fever Clinics were requested to reinforce case finding of pneumonia of unknown etiologies. Fever Clinics, mostly affiliated to the Division of Infectious Diseases in general hospitals, had been established after SARS outbreak in 2003, and since then served as the first line to monitor and manage acute febrile respiratory infections, mostly seasonal influenza in recent years. For 2019 novel coronavirus disease (COVID-19), this time, patients with body temperatures≥37.2°C were asked to firstly visit Fever Clinics, where a triage strategy was implemented ^9^, and probable or possible cases were identified by experts or expert panels, and samples were sent for quick viral detection. On Jan. 21, 2020, the first case of COVID-19 was identified in our Fever Clinic, and up to Feb. 15, 21 cases were diagnosed, with 19 cases confirmed by positive 2019-nCoV results, among 88 patients with pneumonia sent for viral detection. As far as we know, the prevalence of COVID-19 in patients with pneumonia visiting Fever Clinics has not been reported, and there is lack of data comparing the clinical features between COVID-19 and pneumonia of other etiologies.

Therefore, we collected and reviewed the medical records of patients who visited the Fever Clinic in our hospital. We aimed to estimate the prevalence of COVID-19 in pneumonias during this period and to find the unique features of COVID-19 as compared to pneumonias caused by other agents. We believe that the results of our study will add to the knowledge of COVID-19 for practice in Fever Clinics facing newly emerging respiratory infections.

## Methods

### Study design and patient selection

We conducted a retrospective, single-centered study and recruited patients visiting the Fever Clinic at Peking University Third Hospital from January 21 to February 15, 2020. Based on epidemiological history, clinical and radiological manifestations, cases with possible or probably COVID-19 were sent for panel discussion and then for 2019-nCoV detection by RT-PCR. Pediatric patients were not included in our study. The study protocol was approved by the ethics committee of Peking University Third Hospital and the data were analyzed anonymously.

### Diagnostic criteria of COVID-19

The diagnostic criteria of COVID-19 were according to Guidelines for the Diagnosis and Treatment of Novel Coronavirus (2019-nCoV) Infection by the National Health Commission ^10^, which can be accessed in the network http://www.nhc.gov.cn/xcs/zhengcwj/list_gzbd.shtml. Laboratory testing of 2019-nCoV in throat swabs was performed by both Beijing Centers for Disease Control and Prevention (CDC) and Haidian District CDC.

Severity of COVID-19 was evaluated by the following criteria: 1) Mild-moderate: fever and/or respiratory symptoms with pneumonia in radiology examination, without signs of severe or very severe diseases; 2) Severe: presence of one of the following: respiratory rate ≥30 beat/min; SpO_2_ ≤93% at rest; PaO_2_/FiO_2_ ≤300mmHg; 3) Very severe: presence of one of the following: severe respiratory failure requiring mechanical ventilation; shock; complicated with other organ failure and requiring ICU admission.

### Data collection

Demographic and epidemiological data were collected. Epidemiological data included: if a patient came from Wuhan City or Hubei Province in 2 weeks; if a patient ever had contact face-to-face with individuals from Wuhan or Hubei in 2 weeks; if a patient, in 2 weeks, ever had contact face-to-face with individuals who had confirmed COVID-19; if the onset of pneumonia occurred in family members in a short period. Clinical indices and symptoms on presentation were also recorded. Data from laboratory examinations including blood routines, and CT findings were collected. CT features were reviewed by experts of Pulmonary and Critical Care Medicine and Radiology.

### Statistical analysis

Continuous variables were expressed as the mean (standard deviation) or median (25th and 75th percentiles). Categorical variables were expressed as numbers (%). To compare the differences between groups for Continuous variables, *t*-test was used. To compare the differences between groups for categorical variables, chi-square or Fisher exact test was used. Results were considered statistically significant at *P* value <0.05. Statistical analyses were performed using SPSS software, version 22.0 (IBM, Armonk, NY, USA).

## Results

From January 21 to February 15, 2020, based on epidemiological evidence, fever and/or respiratory symptoms, chest radiological findings and blood white blood cell (WBC) results, physicians at the Fever Clinic referred 156 cases to panel discussion by multi-discipline experts. After discussion, 110 were considered to be possible or probable cases of COVID-19, and received 2019-nCoV real-time PCR testing, which was positive in 19 cases (confirmed cases). In another 2 patients, though PCR testing was negative, a clinical diagnosis was made according to epidemiological evidence, consistent clinical and CT findings (clinical cases). These 21 cases were grouped as COVID-19 for this analysis. For the remaining cases with negative viral detection, the diagnosis of COVID-19 was excluded based on inconsistent epidemiological, clinical or radiological data. Among these, 22 were excluded from the analysis because of lack of CT scan or no signs of pneumonia on CT scan, and finally 67 patients with pneumonia of other etiologies were included for analysis as the non-COVID-19 group (see patient selection flowchart in Figure 1).

**Figure 1.**
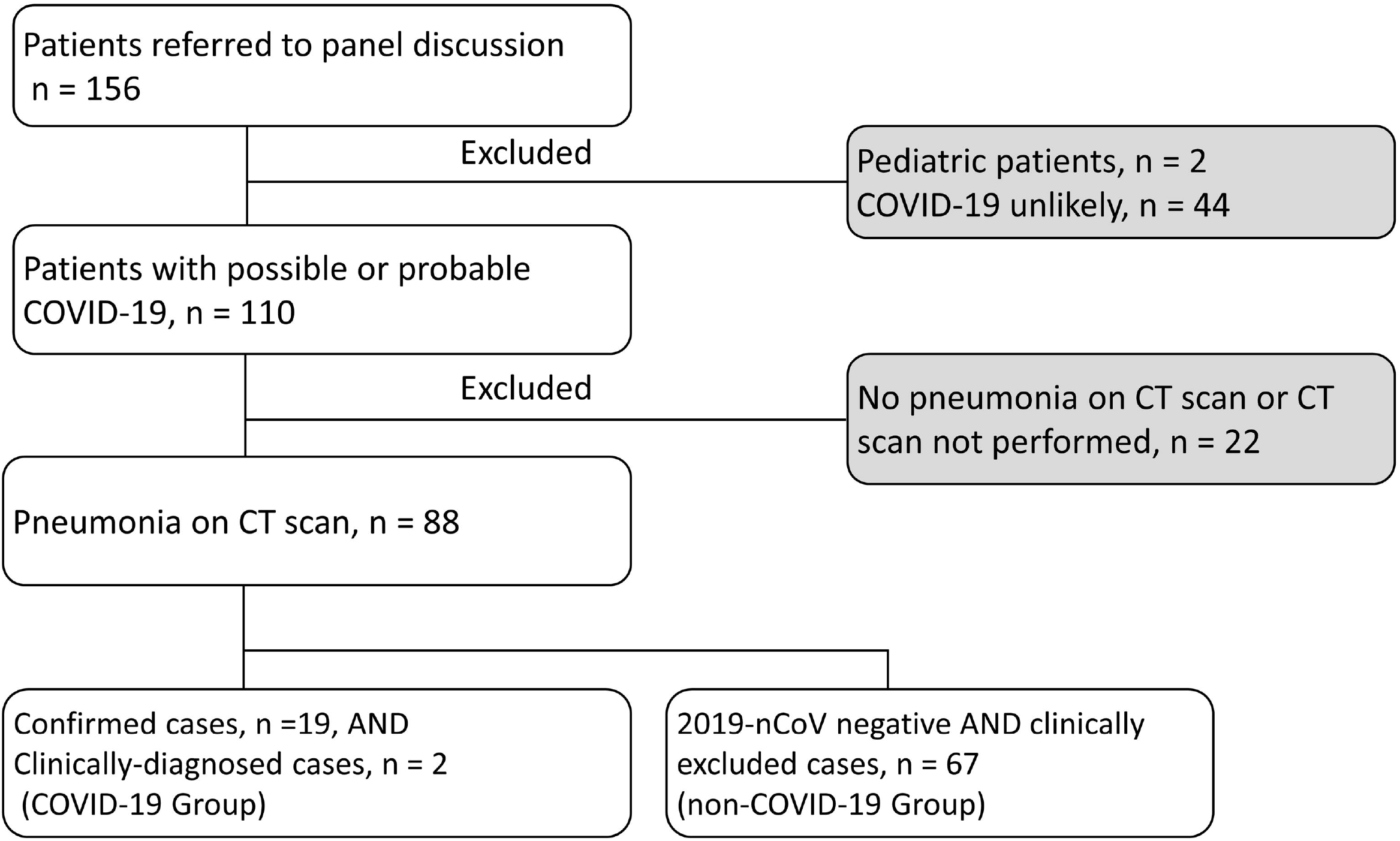
Patient selection diagram. Information of the two patients with clinically-diagnosed COVID-19 (throat swab 2019-nCoV negative): one patient and his wife (2019-nCoV positive) both having fever for more than 3 days and typical features of viral pneumonia on CT scan, had contacted a confirmed case in a same flight. The other patient and his father had fever and cough almost at the same time. Both of them had typical findings of viral pneumonia on CT scans, and 2019-nCoV was detected in the throat swab from his father subsequently. Moreover, his mother had been confirmed to have COVID-19 10 days ago by positive 2019-nCoV testing. COVID-19: 2019 novel coronavirus disease; CT: computed X-ray tomography

In the study period, 2123 patients visited the Fever Clinic because of fever and/or respiratory symptoms, and 342 patients were diagnosed to have pneumonia based on CT scan or in a few cases on Chest X-ray. The prevalence of COVID-19 in all these pneumonia patients was 6.14% (21/342). The prevalence of COVID-19 in cases with CT-confirmed pneumonia sent for 2019-nCoV testing was 23.9% (21/88).

The demographics and baseline characteristics of the 21 patients with COVID-19 were shown in Table 1. The median age was 42.0 years (25th-75th percentile, 34.5-66.0 years). Men and women were equally affected. Nineteen patients (90.5%) had a clear epidemiological history of COVID-19. Seven patients, from 5 family clusters, had close contact with their family members. In the two patients with unclear epidemiological link, one was a social worker contacting a high-flow of people and his wife was confirmed to have COVID-19 by positive 2019-nCoV test about one week later. The other one had no epidemiological history on presentation, but another patient, who had had dinner with him, was also diagnosed to have COVID-19 later. An incubation period was elicited from 12 patients (57.1%), ranging from 2 to 10 days with a median of 6.5 days. Most of the patients were mild-moderate in disease severity (mild-moderate cases, n=18, 85.7%). Three patients were classified as severe cases with SpO_2_ ≤93% on presentation, while no very severe cases were identified in our series.

**Table 1.** Demographics and baseline characteristics of 21 cases of COVID-19

| Demographics and baseline characteristics |  |  |
| --- | --- | --- |
| Age, years |  |  |
| Median (25<sup>th</sup>, 75<sup>th</sup>) |  | 42.0 (34.5, 66.0) |
| Range |  | 24-85 |
| Sex |  |  |
| Male |  | 11 (52.4%) |
| Female |  | 10 (47.6%) |
| Epidemiological history |  |  |
| Imported cases from Wuhan City or Hubei Province |  | 6 (28.6%) |
| Known contact with individuals from Wuhan or Hubei |  | 1 (4.8%) |
| Known contact with cases of confirmed COVID-19 |  | 5 (23.8%) |
| Family aggregation onset |  | 7 (33.3%) |
| None or Unclear |  | 2 (9.5%) |
| Incubation period (day) |  |  |
| Uncertain |  | 9 (42.9%) |
| With clear incubation period |  | 12 (57.1%) |
| Median (25<sup>th</sup>, 75<sup>th</sup>) |  | 6.5 (3.0, 8.0) |
| Range |  | 2-10 |
| Severity of COVID-19 |  |  |
| Mild-Moderate |  | 18 (85.7%) |
| Severe |  | 3 (14.3%) |

Data are presented as n (%) or median (25<sup>th</sup>, 75<sup>th</sup>) or mean (SD). COVID-19: 2019 Novel coronavirus disease; SD: standard deviation;

On presentation, most patients (85.7%) had fever with a mean body temperature of 37.8°C. Cough (42.9%), expectoration (33.3%), fatigue (57.1%), headache or dizziness (38.1%) were common symptoms. Other symptoms included shortness of breath, myalgia or arthralgia, sore throat, nasal symptoms and diarrhea (Table 2). Compared with non-COVID-19 patients, a higher proportion of COVID-19 patients had an identified epidemiological history (90.5% versus 32.8%). Clinical symptoms were similar between cases of COVID-19 and non-COVID-19, except for cough (Table 2).

**Table 2.** Epidemiological history and symptoms on presentation between patients with COVID-19 and non-COVID-19

|  | COVID-19<br>(n = 21) | Non-COVID-19<br>(n = 67) | *P* value |
| --- | --- | --- | --- |
| Epidemiological history |  |  |  |
| Clear | 19 (90.5%) | 41 (32.8%) | < 0.001 |
| Unclear | 1 (4.75%) | 7 (7.5%) |  |
| None | 1 (4.75%) | 40 (59.7%) |  |
| Fever | 18 (85.7%) | 56 (83.6%) | 0.816 |
| Cough | 9 (42.9%) | 53 (79.1%) | 0.001 |
| Expectoration | 7 (33.3%) | 30 (44.8%) | 0.354 |
| Dyspnea or shortness of breath | 1 (4.8%) | 11 (16.4%) | 0.174 |
| Fatigue | 12 (57.1%) | 27 (40.3%) | 0.175 |
| Myalgia or arthralgia | 4 (19.0%) | 17 (25.4%) | 0.553 |
| Sore throat | 2 (9.5%) | 15 (22.4%) | 0.193 |
| Nasal symptoms | 1 (4.8%) | 10 (14.9%) | 0.219 |
| Headache or dizziness | 8 (38.1%) | 15 (22.4%) | 0.153 |
| Diarrhea | 3 (14.3%) | 5 (7.5%) | 0.343 |

Data are presented as n (%).

Blood routine showed that decreased lymphocyte count was present in 38.1% of the COVID-19 patients. More than 80% of COVID-19 patients had normal WBC and neutrophil counts. Compared with COVID-19 patients, the total WBC and neutrophil counts were higher in non-COVID-19 patients. Additionally, more non-COVID-19 patients had higher than normal WBC and neutrophil counts. However, lymphocyte counts and the proportions of patients with decreased lymphocytes were not statistically different between the two groups (Table 3).

**Table 3.** Blood routine test on presentation between COVID-19 and non-COVID-19 groups

|  | COVID-19<br>(n = 21) | Non-COVID-19<br>(n = 67) | *P* value |
| --- | --- | --- | --- |
| Leucocytes ($\times 10^9/L$, normal range 3.5-9.5) | 5.19 (1.47) | 7.21 (2.94) | < 0.001 |
| Decreased | 3 (14.3%) | 4 (6.0%) | 0.047 |
| Normal | 18 (85.7%) | 49 (73.1%) |  |
| Increased | 0 (0.0%) | 14 (20.9%) |  |
| Hemoglobin (g/L) | 145.9 (14.2) | 134.4 (23.4) | 0.036 |
| Platelets ($\times 10^9/L$) | 162.6 (59.0) | 189.8 (67.8) | 0.102 |
| Lymphocytes ($\times 10^9/L$, normal range 1.1-3.2) | 1.26 (0.64) | 1.19 (0.62) | 0.686 |
| Decreased | 8 (38.1%) | 32 (47.8%) | 0.438 |
| Normal | 13 (61.9%) | 35 (52.2%) |  |
| Neutrophils ($\times 10^9/L$, normal range 1.8-6.3) | 3.39 (1.48) | 5.38 (2.85) | < 0.001 |
| Decreased | 3 (14.3%) | 4 (6.0%) | 0.035 |
| Normal | 17 (81.0%) | 42 (62.7%) |  |
| Increased | 1 (4.8%) | 21 (31.3%) |  |

Data are presented as mean (SD) or n (%). SD: standard deviation.

In non-COVID-19 patients, infiltrates on the CT scan mainly involved 1-2 lung lobes, while in COVID-19 patients, lesions involving 4-5 lobes were more common. In COVID-19 patients, lung lesions were mostly peripheral or sub-pleural in distribution, and in severe cases were diffusely distributed (Figure 2). In non-COVID-19 patients, 34.3% showed airway-dominant lesions, while diffuse distribution was found in only 4.5%. The patterns of the lesions showed no significant difference between the two groups. However, centrilobular nodules were observed in non-COVID-19 patients but not in COVID-19 patients (Table 4).

**Table 4.** CT findings between COVID-19 ^a^ and non-COVID-19 patients

|  | COVID-19<br>(n = 20) | Non-COVID-19<br>(n = 67) | *P* value |
| --- | --- | --- | --- |
| Number of lobes involved by pneumonia |  |  |  |
| 1 | 7 (35.0%) | 28 (41.8%) | 0.041 |
| 2 | 3 (15.0%) | 20 (29.9%) |  |
| 3 | 1 (5.0%) | 8 (11.9%) |  |
| 4 | 5 (25.0%) | 3 (4.5%) |  |
| 5 | 4 (20.0%) | 8 (11.9%) |  |
| Involved lobes $\geq 4$ | 9 (45.0%) | 11 (16.4%) | 0.008 |
| Distribution |  |  |  |
| Peripheral or sub-pleural | 16 (80.0%) | 41 (61.2%) | 0.002 |
| airway-dominant | 0 (0.0%) | 23 (34.3%) |  |
| Diffuse | 4 (20.0%) | 3 (4.5%) |  |
| Lesion patterns |  |  |  |
| GGO | 10 (50.0%) | 24 (35.8%) | 0.368 |
| Mixed GGO and consolidation | 9 (45.0%) | 29 (43.3%) |  |
| Consolidation | 1 (5.0%) | 11 (16.4%) |  |
| Centrilobular nodules | 0 (0.0%) | 3 (4.5%) |  |

a: One patient with confirmed COVID-19 showed minimal change on CT scan.
CT: computed X-ray tomography; GGO: Ground-glass opacity; COVID-19: 2019 Novel coronavirus disease

**Figure 2.**
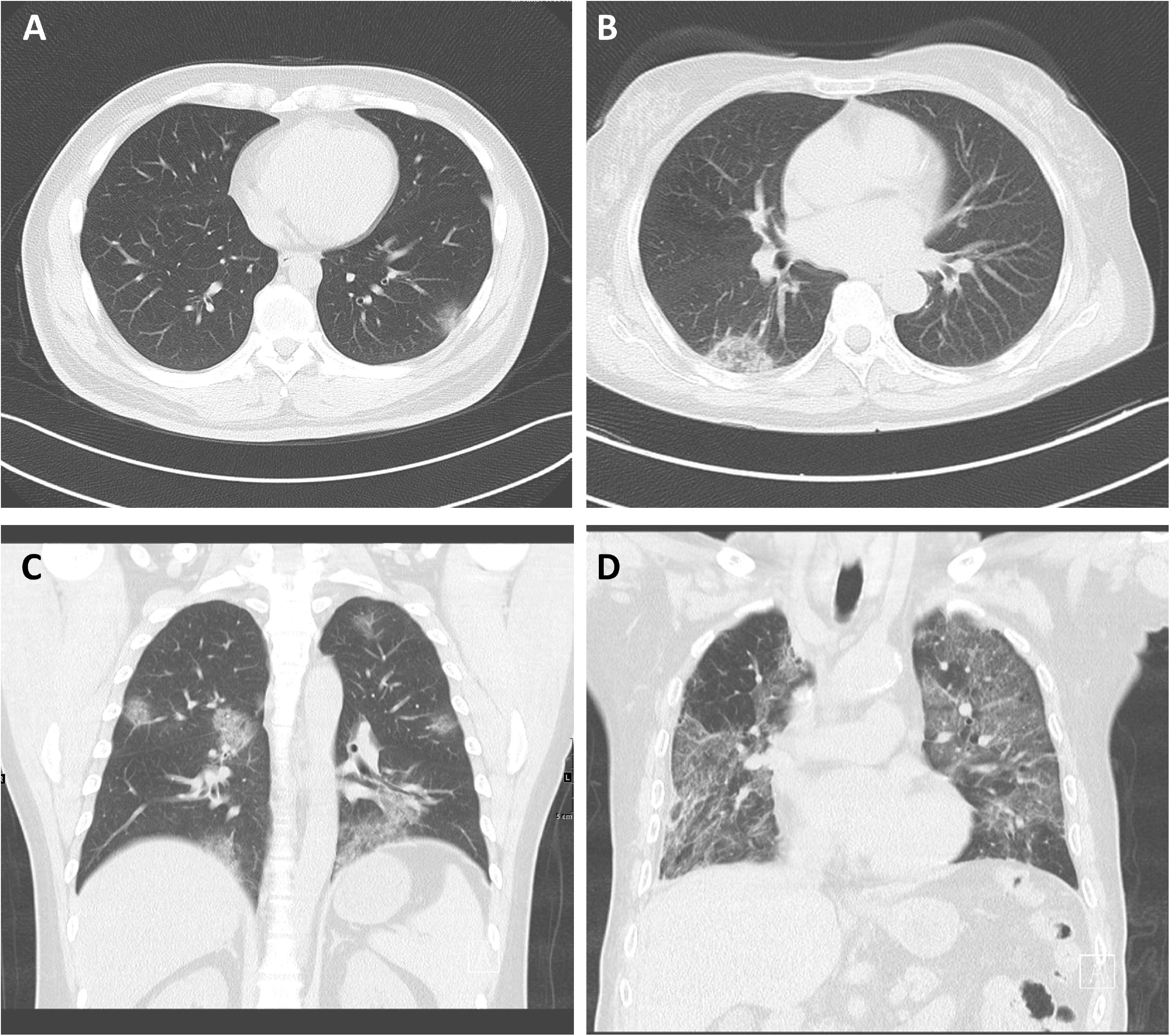
CT findings in 4 patients with confirmed COVID-19. A. Patchy GGO distributed in subpleural area in left lower lung. B. A round-like, mixed GGO and consolidation lesion (with reversed halo sign) in subpleural area in right lower lung. C. Multiple patchy GGO in bilateral lungs, mainly peripheral distribution. D. Diffuse lesions in bilateral lungs (severe case).

## Discussion

As of Feb 15, 2020, 57416 cases of confirmed COVID-19 were reported in 31 provinces, autonomous regions and municipalities in China, among which 49030 were in Hubei Province, while in Beijing municipality, 380 confirmed cases were reported. Since the outbreak in early December, 2019, case finding and reporting of COVID-19 were required for medical staff in Fever Clinics around the country. During this study period, in Beijing, as in other regions of the country, influenza and other respiratory infections were also prevalent; it was a challenge for physicians to identify suspected COVID-19 from patients with fever, particularly from pneumonia of varied etiologies. Our results here provide information on the prevalence of COVID-19 in the busy Fever Clinic of a teaching hospital and differential characteristics from non-COVID-19 pneumonias.

From January 21, 2019, when the first case of COVID-19 was identified, up to Feb.15, 2020, 2123 patients visited our Fever Clinic because of fever and/or respiratory symptoms, and 342 patients were confirmed to have pneumonia by CT scan or in a few cases by Chest X-ray. The prevalence of COVID-19 in all these pneumonia patients was 6.14% (21/342). The prevalence of COVID-19 in cases with pneumonia sent for 2019-nCoV testing was 23.9% (21/88).

We found that epidemiological history was extremely important for identification of suspected cases of COVID-19 and for differential diagnosis. Over 90% of the COVID-19 patients had clearly identified epidemiological evidence, while in non-COVID-19 patients, epidemiological links could only be found in 32.8%. In our series, most cases were imported from the city of Wuhan or Hubei Province, including traveling to or living in Wuhan or other cities in Hubei, and a few had a history of close contact with individuals coming from the above areas. In the later phase of our study, contact with family members with confirmed COVID-19 was an epidemiological feature and five family clusters were found. A report from Chinese CDC showed that the incidence of COVID-19 outside Hubei Province peaked in Jan 24-27 and most cases (68.6%) had ever exposed to Wuhan or Hubei within 14 days ^11^. Weight of exposure history related to Wuhan and Hubei in confirmed cases was decreasing after February ^11^. Consistent to previous reports, the incubation period of our cases was in a range of 2-10 days, with a median of 6.5 days ^12-14^.

A unique feature of our study was the comparison between COVID-19 and non-COVID-19 cases visiting the same Fever Clinic at a certain period, with the finding that the total WBC count and the neutrophil count were different between the two groups of patients. Increased total WBC and neutrophil counts were more common in patients with other causes of pneumonia. In contrast, although decrease of lymphocytes was also common in our study, the difference between COVID-19 and non-COVID-19 cases was not significant, probably due to the fact that pneumonias enrolled for this analysis were mostly viral(for example, influenza) or atypical, as classical bacterial pneumonia had been excluded by first screening/triage. Additionally, the percentage of our patients with decreased lymphocytes was not as high as that reported in some previous studies ^2,15^, perhaps due to differences in disease severity. Blood lymphopenia was observed in up to 85% of critically ill patients with COVID-19 ^16^. Compared with mild and moderate cases, severe cases were more likely to have lymphopenia ^2,16,17^. A recent paper by Xu and colleagues showed a lymphocyte decrease in 38% of the cases within symptom onset ≤10 days ^18^, which was consistent with our data. Therefore, we speculate that persisting decrease of lymphocytes may be an indicator of severe disease, rather than a criterion for the diagnosis of COVID-19.

Unlike pneumonia caused by other pathogens, COVID-19 has some unique characteristics on the CT scan. Firstly, multi-lobe involvement was more common in COVID-19 as compared to non-COVID-19 pneumonia. The lesions, including GGO, mixed GGO and consolidation, or patchy consolidation, were mainly distributed in peripheral and sub-pleural areas in mild to moderate cases, while in severe cases diffuse ground glass lesions were present. In the study by Pan and colleagues, 11.1% and 44.4% of the 63 patients showed four and five lobe involvement, with patchy/punctate GGO and patchy consolidation being the dominant patterns on CT scan ^19^. Other studies also demonstrated similar CT findings ^20^.

Our study had several limitations. As a single-center, retrospective study of patients mainly from Haidian District in Beijing, the prevalence of COVID-19 in this Fever Clinic was not representative. The sample was small-sized and mostly mild and moderate in severity, which may not reflect the whole picture of the disease. Furthermore, the pathogens causing pneumonia in patients without COVID-19 were not identified, therefore we were unable to evaluate the prevalence of other common viral pneumonias of this season, such as influenza pneumonia and adenoviral pneumonia, making specific comparisons between different viral pneumonias unavailable.

## Conclusions

During the study period when COVID-19 was epidemic, the prevalence of COVID-19 in patients with pneumonia visiting our Fever Clinic was 6.14%. Most of the COVID-19 cases were mild and moderate in severity and more than 90% had identified epidemiological exposure. Changes of WBC and neutrophil counts showed clinical significance for differential diagnosis of pneumonia caused by 2019-nCoV or other pathogens. CT findings of COVID-19 were characteristic, including the number of affected lobes, lesion patterns and distribution, which may be helpful for clinicians to identify suspected cases of COVID-19 from pneumonia of other etiologies.

## Data Availability

The data used to support the findings of this study are available from the corresponding author upon request.

